# Effects of a 24-week resistance exercise program on Alzheimer’s disease brain signatures in cognitively normal older adults: results from the AGUEDA trial

**DOI:** 10.1101/2025.07.23.25332047

**Authors:** Javier Sanchez-Martinez, Patricio Solis-Urra, Beatriz Fernandez-Gamez, Javier Fernández-Ortega, Lucía Sánchez-Aranda, Kirk I. Erickson, Francisco B. Ortega, Irene Esteban-Cornejo

## Abstract

**Objective:** To examine the effects of a 24-week resistance exercise (RE) program on Alzheimer’s disease (AD) brain signatures in cognitively normal older adults; and to identify moderating factors, assess the association between AD brain signatures changes and cognitive outcomes, including mediation by AD brain signatures

**Methods:** Ninety participants (72±4 years, 58% female) were randomized to a RE group (3 sessions/week, 60 min/session, n=46) or a wait-list control group (CG, n=44). T1- and diffusion-weighted MRI (3T) were acquired pre- and post-intervention. AD thickness/volume and gray matter mean diffusivity (GMMD) signatures were derived from cortical and hippocampal regions. Baseline adjusted mixed models tested intervention effects. Moderators included age, sex, education, APOE4 status, amyloid beta (Aβ) status, and baseline AD signatures. Associations with cognition and possible mediators were explored.

**Results:** The RE group showed reduction in the AD thickness/volume signature (−0.23 standardized mean difference [SMD]; 95% CI, −0.43 to −0.02), but no effect on the AD GMMD signature (0.08 SMD; 95% CI, −0.13 to 0.29) vs CG. Aβ status moderated the effect, with Aβ-positive participants in the RE group showing a larger reduction in AD thickness/volume signature compared to the CG (−0.64 SMD; 95% CI, −1.09 to −0.18), whereas no effect was found in Aβ-negative participants. AD thickness/volume signature reductions were associated with better executive function, and AD GMMD signature reductions with attentional/inhibitory control improvements. Changes in AD signatures did not mediate changes in cognition.

**Conclusion:** A 24-week RE intervention reduced AD thickness/volume signature, especially in Aβ-positive participants, and was linked to improved executive function. No effects were observed on the AD GMMD signature. These findings suggest RE may support brain health by delaying structural changes characteristic of preclinical stages of AD. Further research is needed to replicate and confirm these results.

## INTRODUCTION

Brain imaging markers hold promise for the early detection of cognitive impairment, and conversion to diagnosed dementia cognitive.^1^ Cortical thickness is a key predictor of Alzheimer’s disease (AD) progression,^2^ with higher AD-signature based on cortical thickness linked to a lower risk of dementia in both cognitively healthy adults and those with mild cognitive impairment.^2–4^ In contrast, elevated gray matter mean diffusivity (GMMD) in AD-related regions is associated with increased AD risk.^3^ Additionally, reduced cortical thickness has been observed in amyloid beta (Aβ) positive individuals, independent of apolipoprotein E *ε*4 (APOE4) status.^5^ Moreover, a longitudinal study found greater increases in cortical mean diffusivity over time (with a follow-up ranging from 2 to 8 years) in early stages of AD, with the highest increase in Aβ-positive/tau-positive participants, followed by Aβ-positive/tau-negative participants.^6^ These findings highlight the predictive potential of brain imaging markers in AD and indicate the complexity of brain biomarkers, such that some brain imaging changes could precede the onset of others. This complexity underscores the need for more multi-modal imaging studies to better understand these issues.

Although evidence suggests that exercise affects brain structure in cognitively unimpaired older adults, there remains significant heterogeneity across the field. Some studies have reported increases in total brain volume,^7^ hippocampal volume,^8,9^ and cortical thickness^10^ after exercise interventions. However, others studies have found no effects of exercise on cortical thickness, gray matter, or hippocampal volume.^10–13^ Research examining the effect of exercise on AD-related brain signatures is still limited. For instance, a two-year multimodal intervention (including diet, exercise, cognitive training, and vascular risk monitoring) in older adults at increased risk for dementia did not impact AD-signature cortical thickness.^14^ Another study reported that a three-month supervised walking intervention increased GMMD, which correlated with cognitive improvements.^15^ However, to our knowledge, no prior studies have examined the effects of resistance exercise (RE) on AD brain signatures. Given their significance as early biomarkers, it is relevant to investigate how different exercise modalities, such as RE, influence AD-related structural and microstructural brain changes and their potential role in preventing or delaying the onset of dementia and AD.

The primary aim of this study was to investigate the effects of a RE program on AD thickness/volume and GMMD signatures in cognitively normal older adults. The secondary aim was to identify potential moderating factors and examine the associations between changes in AD brain signatures and cognitive outcomes, including the potential mediating role of AD brain signatures in the effect of RE on cognitive outcomes. We hypothesized that: (i) the RE program would increase AD thickness/volume and reduce GMMD signatures; (ii) changes in AD brain signatures would be associated with cognitive function; and (iii) AD brain signatures would mediate the effect of RE exercise group allocation on cognitive outcomes.

## METHODS

### Design and participants

Our study was conducted under the framework of the AGUEDA trial (“Active Gains in brain Using Exercise During Aging”).^16^ AGUEDA is a single-site, two-arm, single-blind randomized controlled trial (RCT). Participants were recruited from community-dwelling older adults in the city of Granada (Spain) and surrounding areas from March 2021 to May 2022. Details of the inclusion and exclusion criteria were previously published in the study protocol.^16^ Ninety cognitively normal older adults (65-80 y) were randomly assigned to a 24-week RE group (n = 46) or a wait-list control group (CG, n = 44). The study followed the principles of the Declaration of Helsinki and was approved by the Research Ethics Board of the Andalusian Health Service (CEIM/CEI Provincial de Granada; #2317-N-19). The trial was registered on Clinicaltrials.gov (ClinicalTrials.gov Identifier: NCT05186090). All participants provided written informed consent once all study details were explained. Further methodological details, including statistical analysis plan, sample size calculation, randomization, blinding, participant flow, intervention and comparator delivery, and adverse events, have been previously described.^16,17^ Protocols are deposited at GitHub (https://github.com/aguedaprojectugr). The results are reported in accordance with the Consolidated Standards of Reporting Trials (CONSORT) statement (Supplementary Table 1).^18^

**Table 1.** Baseline characteristics for the overall sample and allocated groups.

|  | Overall | RE | CG |
| --- | --- | --- | --- |
| n | 90 | 46 | 44 |
| Age (y) | 71.75 (3.96) | 71.90 (4.21) | 71.58 (3.73) |
| Age group, older (%) | 40 (44.4) | 20 (43.5) | 20 (45.5) |
| Sex, female (%) | 52 (57.8) | 27 (58.7) | 25 (56.8) |
| Body mass index (kg/m<sup>2</sup>) | 28.50 (4.23) | 28.35 (4.43) | 28.67 (4.05) |
| Education level (y) | 11.54 (4.90) | 11.17 (5.26) | 11.93 (4.53) |
| Education level, high category (%) | 33 (36.7) | 14 (30.4) | 19 (43.2) |
| *Biomarkers* |  |  |  |
| APOE4 status, positive carrier (%) | 13 (14.8) | 7 (15.6) | 6 (14.0) |
| PET Aβ (centiloid) | 6.89 (25.16) | 3.60 (23.33) | 10.33 (26.78) |
| PET Aβ status, positive (%) | 19 (21.1) | 8 (17.4) | 11 (25.0) |
| pTau-217 (pg/ml) | 0.30 (0.15) | 0.31 (0.16) | 0.30 (0.13) |
| *AD brain signatures (z-scores)* |  |  |  |
| AD thickness/volume signature | 0.00 (1.00) | 0.19 (0.89) | -0.20 (1.08) |
| AD gray matter mean diffusivity signature | 0.00 (1.00) | -0.01 (1.07)<sup>a</sup> | 0.01 (0.93)<sup>b</sup> |
| *Cognitive function (z-scores)* |  |  |  |
| Attentional/inhibitory control | -0.01 (1.00) | 0.04 (0.98) | -0.06 (1.03) |
| Episodic memory | 0.00 (1.01) | -0.01 (0.85) | 0.01 (1.15) |
| Executive function | -0.01 (1.00) | -0.03 (1.07) | 0.01 (0.94) |
| Processing speed | -0.01 (1.00) | 0.04 (0.85) | -0.06 (1.15) |
| Visuospatial processing | -0.01 (1.00) | -0.09 (0.94) | 0.08 (1.06) |
| Working memory | -0.02 (0.99) | 0.01 (1.05) | -0.04 (0.94) |

Data showed as mean (standard deviation). Abbreviations: AD, Alzheimer's disease; APOE4, apolipoprotein E $\epsilon$ 4 allele; CG, wait-list control group; PET, Positron Emission Tomography; RE, resistance exercise group; y, years.
<sup>a</sup> n = 45.
<sup>b</sup> n = 41.

### Intervention

The 24-week RE program consisted of 3 sessions/week. Each session lasted 60 min and included a combination of upper and lower limb exercises using elastic bands and the participant’s own body weight (3 sets of 8 exercises). The target-intensity was 7-8 out of 10 of Borg’s Rating of Perceived Exertion.^19^ See Fernandez-Gamez 2023^20^ for more details of the RE program implemented in the AGUEDA trial based on the CERT (Consensus on Exercise Reporting Template). Participants assigned to the wait-list CG could perform the exercise program after finishing the post-evaluations.

### Image acquisition

Magnetic resonance imaging (MRI) was performed on a 3T scanner (Siemens Magnetom PRISMA Fit) equipped with a 64-channel head coil at the Mind, Brain and Behavior Research Centre (CIMCYC), University of Granada. T1-weighted magnetization-prepared rapid acquisition gradient echo (MPRAGE) sequences were acquired for structural brain scans (parameters: sagittal, 0.8 mm isotropic resolution, echo/inversion/repetition times = 2.31/1060/2400 ms, field of view = 256 mm, 224 slices; acquisition time: 6 min 38 seconds). Diffusion-weighted images were acquired using a diffusion sequence (parameters: Resolution: 2 × 2 × 2 mm, TE/TR = 95.6/2800 ms, multiband factor = 4, b-values of 1500, 3000 s/mm^2^, 64 gradient directions).

### Structural image preprocessing

T1-weighted MPRAGE images were processed using FreeSurfer 7.4.1 (https://surfer.nmr.mgh.harvard.edu) on Neurodesk (https://www.neurodesk.org/) with the following three steps for longitudinal processing. The pipeline is summarized in Supplementary Figure 1. First, in the cross-sectional processing, the two images (pre- and post-intervention) of each subject were independently processed using the “recon-all” pipeline, with the −3T flag (*recon-all −3T)*. Second, a within-subject template (*recon-all-base*) was created by averaging the processed images from the cross-sectional processing. Third, longitudinal processing used the outputs of the previous steps to obtain the final images (*recon-all-long*). https://surfer.nmr.mgh.harvard.edu/fswiki/FsTutorial/LongitudinalTutorial.

### Structural image quality control

Quality control (QC) was independently performed by two researchers (JSM, JOF) following the ENIGMA Consortium Cortical QC Protocol 2.0 (https://enigma.ini.usc.edu). Image quality of the FreeSurfer parcellation output was categorized as *pass, moderate* or *fail*, following ENIGMA protocol recommendations (https://enigma.ini.usc.edu/protocols/imaging-protocols/). See Results for a summary of QC results.

### Diffusion-weighted image preprocessing

Diffusion-weighted images (DWI) were processed on Neurodesk (https://www.neurodesk.org/) using command modules from MRtrix3 v3.04,^21^ FSL v6.0.7.16, and ANTs v2.6.0. The pipeline is summarized in Supplementary Figure 2. First, the diffusion-weighted gradient scheme (bvecs/bval files) was imported during the conversion of raw NifTi images to MRtrix3 format (.mif). The preprocessing steps included denoising (*dwidenoise* command), and removal of Gibbs ringing artifacts (*mrdegibbs* command). Motion and distortion correction with FSL’s eddy and topup tools within the *dwifslpreproc* script in MRtrix3, with phase encoding set to *-pe_dir j*, and including - eddy_options (“--repol^22^ --cnr_maps --slm=linear) Images acquired with a different phase encoding (n = 3) were excluded. Bias field correction was applied using the ANTs algorithm via the *dwibiascorrect* command in MRtrix3, and brain masking was done with SynthStrip (*mri_synthstrip*, v7.4.1).^23^ Diffusion tensors were computed using *dwi2tensor*, and mean diffusivity (MD) was calculated as the average of the three eigenvalues (λ1, λ2, λ3) at each voxel. To avoid physically implausible values, negative eigenvalues were set to zero prior to computation, as described in previous studies.^24,25^

### DWI quality control

Two researchers (JSM, MTRP) independently assessed raw image quality using a 4-point scale:^26^ 1= “excellent”, 2 = “minor”, 3 = “moderate, and 4 = “severe”, based on motion and artifacts (e.g., spiking, ghosting, missing slices). Images rated as severe were excluded; those rated as moderate were included in the main analyses but excluded from sensitivity analyses. Automatic QC was performed using EDDY QC^27^ (*-eddyqc_all* in *dwifslpreproc*, MRtrix3). Extracted metrics included average absolute motion, average relative motion, outliers’ percentage, and average contrast-to-noise-ratio (CNR). Signal-to-noise-ratio (SNR) in the b0 was computed using the denoised image, the residual noise from denoising, and the brain mask generated by SynthStrip. Images exceeding thresholds (absolute motion ≥2 mm, relative motion ≥0.5 mm, outliers ≥2%, SNR ≤20, CNR ≤1.5)^28^ in two or more metrics were excluded. See Results for a summary of QC results.

### Gray matter mean diffusivity

First, the b0 image (extracted from the preprocessed DWI) was registered to its respective T1 image (using the T1.mgz image from the structural longitudinal processing) with FSL’s *epi_reg* command,^29^ which performs boundary-based registration. The resulting transformation matrix was then applied to the MD images using the *FLIRT* command. A spatial smoothing with a 6-mm full-width at half maximum (FWHM) Gaussian kernel was applied to the MD images using the *mrfilter* command.

To limit the analysis to gray matter voxels and minimize the potential impact of partial volume effects, individual masks for gray matter and cerebrospinal fluid (CSF) were generated. The gray matter mask was created by processing each *aparc+aseg*.*mgz* file with the *5ttgen freesurfer* command in MRtrix3, which produced binary segmentation images of cortical gray matter, sub-cortical gray matter, white matter, and CSF. The cortical and subcortical gray matter images were extracted, merged into a single image, and then resampled to match the resolution and voxel grid of the corresponding FreeSurfer T1 image using the *antsApplyTransforms* command with nearest-neighbor interpolation (*--interpolation NearestNeighbor*) and an identity transformation *(−t identity*), ensuring no geometric transformation was applied.

As recommended,^30^ partial volume effects arising from CSF contamination must be addressed when analyzing GMMD. A CSF mask was created from the preprocessed DWI using multi-tissue constrained spherical deconvolution in MRtrix3, executed with the *dwi2fod msmt_csd* command for multi-shell diffusion data. This approach estimated the contribution of white matter-like, gray matter-like and free water CSF-like signals within each voxel. The three tissue compartment images were normalized using the *mtnormalise* command. The first volume of the white matter compartment was extracted using *mrconvert*, then summed with the gray matter and CSF signals using *mrcalc*. An image reflecting the proportion of CSF signal in each voxel was generated using *mrcalc*, and the b0-to-T1 transformation matrix was applied using the FLIRT command. Following prior research and recommendations,^15,31^ each subject’s CSF mask was binarized with a threshold of 0.5 (including voxels with at least 50% CSF-like signal) using *mrcalc*. The final individual gray matter mask was generated by excluding any voxels overlapping with the CSF mask. This gray matter mask was then applied to the MD image to create a filtered GMMD image, which contains MD values only in gray matter voxels (Supplementary Figure 2).

### Processing of Desikan-Killiany Atlas for Gray Matter Mean Diffusivity

Regions from the Desikan-Killiany parcellation atlas were created for each subject using the *labelconvert* command in MRtrix3 on FreeSurfer’s *aparc+aseg*.*mgz* image. Individual masks for each region of interest (ROI) were generated using the *mrcalc* command. Finally, the mean GMMD for each ROI was calculated by averaging the GMMD values of all voxels within the ROI using the *mrstats* command.

### Main outcomes: Alzheimer’s disease brain signatures

AD thickness/volume and GMMD signatures were computed following Williams et al. (2021) methodology.^3^ The AD brain signatures comprise a weighted average of cortical thickness in seven ROIs (entorhinal cortex, middle temporal gyrus, bank of superior temporal sulcus, superior temporal gyrus, isthmus cingulate, lateral orbitofrontal cortex, and medial orbitofrontal cortex), along with hippocampal volume (**Figure 1**). This composite metric is referred to as the “thickness/volume signature”. The weights assigned to each ROI for the left and right hemispheres are detailed in Williams et al. (2021). In accordance with this methodology, structural and diffusion data for each ROI were adjusted for age, and hippocampal volume was further regressed for estimated intracranial volume to account for differences in head size. Standardized residuals for each ROI were computed separately at each time point (baseline and post assessment), then weighted and summed to calculate the thickness/volume signature scores. The same weights used for structural data were applied to the GMMD values of each ROI, including the hippocampus.

**Figure 1.**
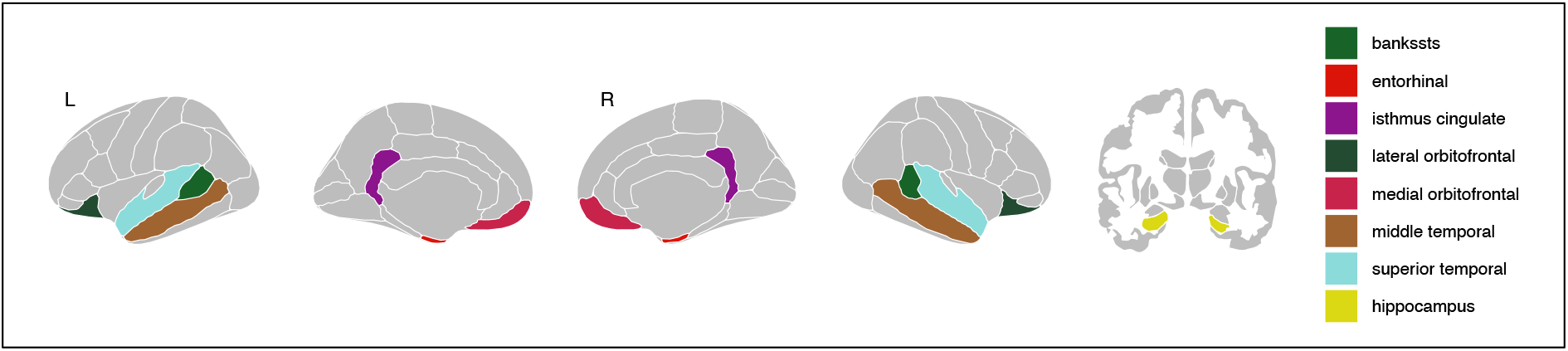
Brain regions used to compute Alzheimer’s disease brain signatures. Abbreviation: bankssts, bank of superior temporal sulcus.

Cortical thicknesses for each ROI were extracted from FreeSurfer’s outputs files (*lh*.*aparc*.*stats* and *rh*.*aparc*.*stats* files), while hippocampal volumes for the left and right hemispheres, as well as the estimated intracranial volume, were obtained from the *aseg*.*stats* file. All files contain the Desikan-Killiany atlas parcellation.^32^

### Cognitive function

At baseline and after the intervention, participants completed a comprehensive neuropsychological evaluation that measures different cognitive domains (Supplementary Table 2). Thus, attentional/inhibitory control, episodic memory, processing speed, visuospatial processing, working memory and executive function scores were used based on a previous analysis of this sample.^17^

### Statistical analysis

#### Main analysis

All statistical analyses were performed using R Statistical Software (Version 4.4.1). We used constrained longitudinal mixed models (baseline adjusted), applying the “LMMstar” and “lme4” packages,^33,34^ with restricted maximum likelihood estimation, to test the effects of the intervention on AD thickness/volume and GMMD signatures. The models included time and group as a categorical fixed effect, and group-by-time interaction, with the intercept specified as a random effect. Unequal variance was allowed across time and group. Statistical significance was set at *p* <0.05 without adjustments for multiple comparisons. Estimated marginal means, within-group differences, and between-group differences were calculated by “*emmeans”* package.^35^ An intention-to-treat approach was used as the primary analysis, while per-protocol analysis (≥ 80% session attendance) as secondary/exploratory. Main intervention effects are presented as z-scores, which represent standardized mean differences (SMDs), indicating how post-exercise values deviate from baseline mean and standard deviation (SD). Effect sizes were interpreted as follows: ~0.2 SDs for a small effect, ~0.5 SDs for a medium effect, and ~0.8 SDs for a large effect. Between-group z-score differences were computed as the exercise group minus control group. Significance was set at *p* <0.05.

#### Sensitivity analysis

For the AD thickness/volume signature, a sensitivity analysis was conducted by excluding images with parcellation issues determined by the visual inspection (ENIGMA protocol) in the ROIs used to compute the signature. Additionally, we examined the effects of the intervention on AD signatures derived from cortical thickness and volumes, incorporating alternative methodologies that included additional brain regions (see Supplementary Figure 3).^36–41^ For the AD GMMD signature, a sensitivity analysis was performed by excluding low-quality images identified through visual QC, automated QC, and those with incorrect acquisition parameters. See Results for a summary of QC results.

#### Moderator effects

Effect moderation was examined by treating age, sex (male, female), education level, APOE4 carrier status, baseline Aβ burden, baseline pTau217 levels, baseline AD thickness/volume signature level, and baseline AD GMMD signature level as interaction terms. Age was categorized as younger (≤72 years) and older (>72 years). Education level was classified as low (≤12 years of education) and high (>12 years of education). Participants with ≥1APOE4 alleles were considered positive carriers. Brain Aβ deposition was assessed before and after the intervention using a PET scan with a Biograph-Vision 600 Edge Positron Emission Tomography/Computed Tomography (PET/CT) digital scanner (Siemens, Erlangen, Germany) at Virgen de las Nieves University Hospital in Granada. Aβ positivity was determined using the centiloid method, with a cut-off of >12, quantified by [18F]Forbetaben amyloid-PET scan.^17^ PTau-217 levels were measured using a Single molecule array (SIMOA) method on an HD-X instrument with commercial assays from Quanterix. PTau-217 were further categorized as high or low based on the median. AD thickness/volume signature and AD GMMD signature levels were categorized as high or low based on the median. Significance was set at *p* <0.05.

#### Associations of changes

Linear regression models were used to examine the associations between z-score changes in AD thickness/volume and GMMD signatures (predictors, analyzed separately) and z-score changes in cognitive domains (dependent variables). Statistical significance was set at *p* <0.05.

#### Mediation analysis

We conducted a mediation analysis (“lme4” and “mediation” packages)^34,42^ to assess whether changes in the z-score AD thickness/volume or z-score AD GMMD signature mediate the relationship between exercise or control conditions and cognitive outcomes (z-score). We included the outcome of interest at baseline as a covariate. We report total effects (overall impact of the predictor variable X on the outcome variable Y), direct effects (effect of X on Y without considering the mediator M), and indirect effects (mediation effect, calculated as the total effect minus the direct effect). The estimated indirect effect reflects the impact of X on Y through the mediator M. Significance was set at *p* <0.05.

## RESULTS

Table 1 describes the baseline characteristics of the sample.

### Quality control

Details on participant inclusion and exclusion based on brain image quality are summarized in Supplementary table 3. After applying the ENIGMA protocol, most image parcellations were rated as “pass” (64.7%) or “moderate” (35.3%), with none rated as “fail”. Due to parcellation issues of ROIs in the “moderate” category, 21 participants’ images (Pre = 11; Post = 10) were excluded from the sensitivity analysis of the AD thickness/volume signature. For visual DWI QC, 36.1% participant’s images were rated as “excellent”, 55% as “minor”, 8.3% (14 images) as “moderate”, and 0.6% (1 image) as “severe”. Automatic QC identified 2 low-quality images. Additionally, 3 images were obtained using an incorrect phase direction. Thus, 5 participants’ images were excluded from the main analysis (Pre = 4, Post = 1), and an additional 29 participants’ images (Pre = 11, Post = 18) were excluded from the sensitivity analysis of the AD GMMD signature.

### Effects on main outcomes: AD thickness/volume and GMMD signatures

A significant group × time effect was found for the AD thickness/volume signature (Figure 2A), indicating a significant reduction in the RE group compared to the CG with a small effect size (SMD: −0.23 [95%CI, −0.43 to −0.02]; p = 0.032). The per-protocol analysis yielded a consistent result (SMD: −0.22 [95%CI, −0.43 to −0.01]; p = 0.039). Consistent results were found using alternative methodologies to compute the AD signature using cortical thickness or volume, and for the sensitivity analysis (Supplementary Table 4). No group x time effect was found for AD GMMD signature (SMD: 0.08 [95%CI, −0.13 to 0.29]; p = 0.457) (Figure 2C). Per-protocol analysis (SMD: 0.08 [95%CI, −0.14 to 0.29]; p = 0.474) and sensitivity yielded a consistent result (Supplementary Table 5).

**Figure 2.**
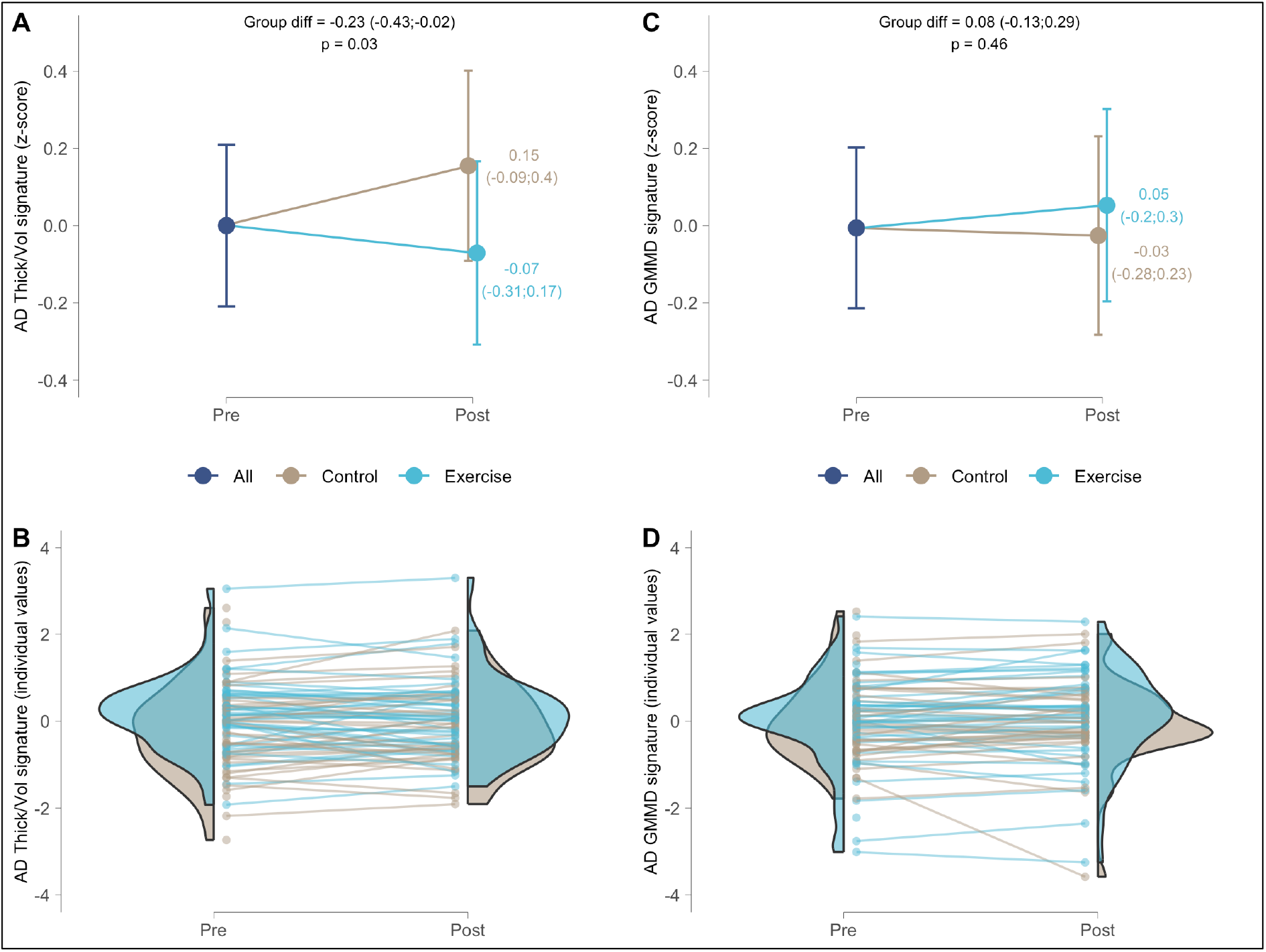
Effects of a 24-week resistance exercise program on Alzheimer’s disease brain signatures in older adults. (A) Estimated marginal means at each time point (and 95% confidence intervals) for AD thickness/volume signature (z-score). (B) Individual z-scores values at each time point for AD thickness/volume signature. (C) Estimated marginal means at each time point (and 95% confidence intervals) for AD GMMD signature (z-score). (D) Individual z-scores values at each time point for AD GMMD signature. Abbreviations: AD, Alzheimer’s disease; GMMD, gray matter mean diffusivity; Thick/Vol, thickness/volume.

### Moderation effects

For the AD thickness/volume signature, we found a moderation effect for Aβ burden (Figure 3A) in both intention-to-treat (p=0.034) and per-protocol analyses (p=0.038). In Aβ-positive older adults, we found a reduction of AD thickness/volume signature in the RE group compared to the CG with a large effect size (SMD: −0.64 [95%CI, −1.09 to −0.18]; p = 0.010) (Figure 3B). In Aβ-negative participants, no group × time effect was found in the AD thickness/volume signature (SMD: −0.10 [95%CI, −0.33 to 0.13]; p = 0.394) (Figure 3C). For AD GMMD signature, there were no moderation effects (Figure 3F).

**Figure 3.**
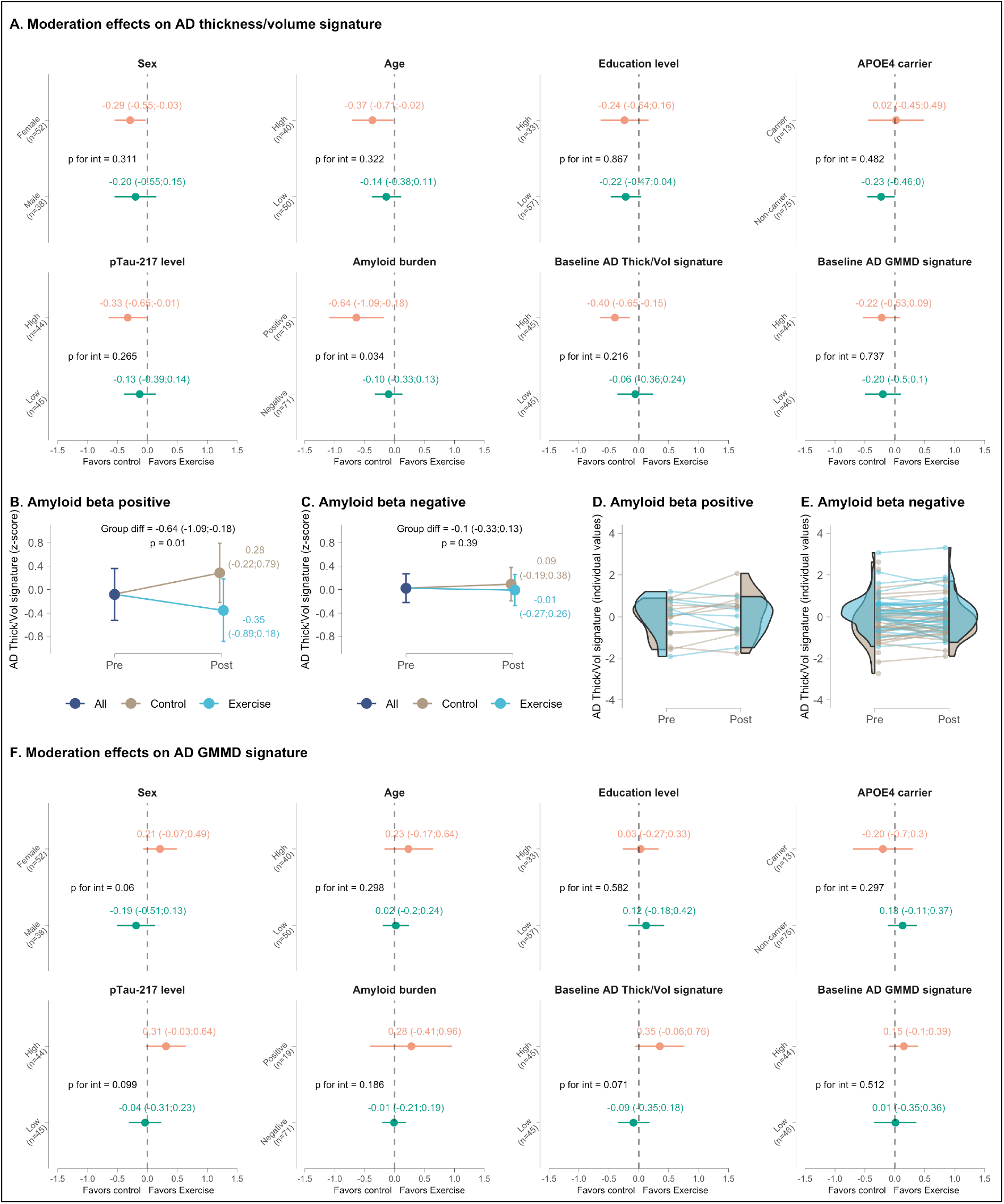
Moderator analyses of the effect of a 24-week resistance exercise program on Alzheimer’s disease (AD) brain signatures in older adults. (A) Effect of moderators on AD thickness/volume signature, with a significance found for amyloid beta (Aβ) burden. (B) Estimated marginal means at each time point (and 95% confidence intervals) for AD thickness/volume signature in Aβ-negative participants. (C) Estimated marginal means at each time point (and 95% confidence intervals) for AD gray matter mean diffusivity signature in Aβ-negative participants. (D) Individual z-scores values at each time point for AD thickness/volume signature in Aβ-positive participants. (E) Individual z-scores values at each time point for AD thickness/volume signature in Aβ-negative participants. (F) No effect of moderators on AD gray matter mean diffusivity signature. Abbreviations: AD, Alzheimer’s disease; APOE4, apolipoprotein E *ε*4, GMMD, gray matter mean diffusivity; Thick/Vol, thickness/volume.

### Associations between changes in AD brain signatures and cognitive domains

Changes in the AD thickness/volume signature were negatively associated with executive function, and particularly in the exercise group (Figure 4A). Regarding the AD GMMD signature, a negative association was identified with attentional/inhibitory control (Figure 4B).

**Figure 4.**
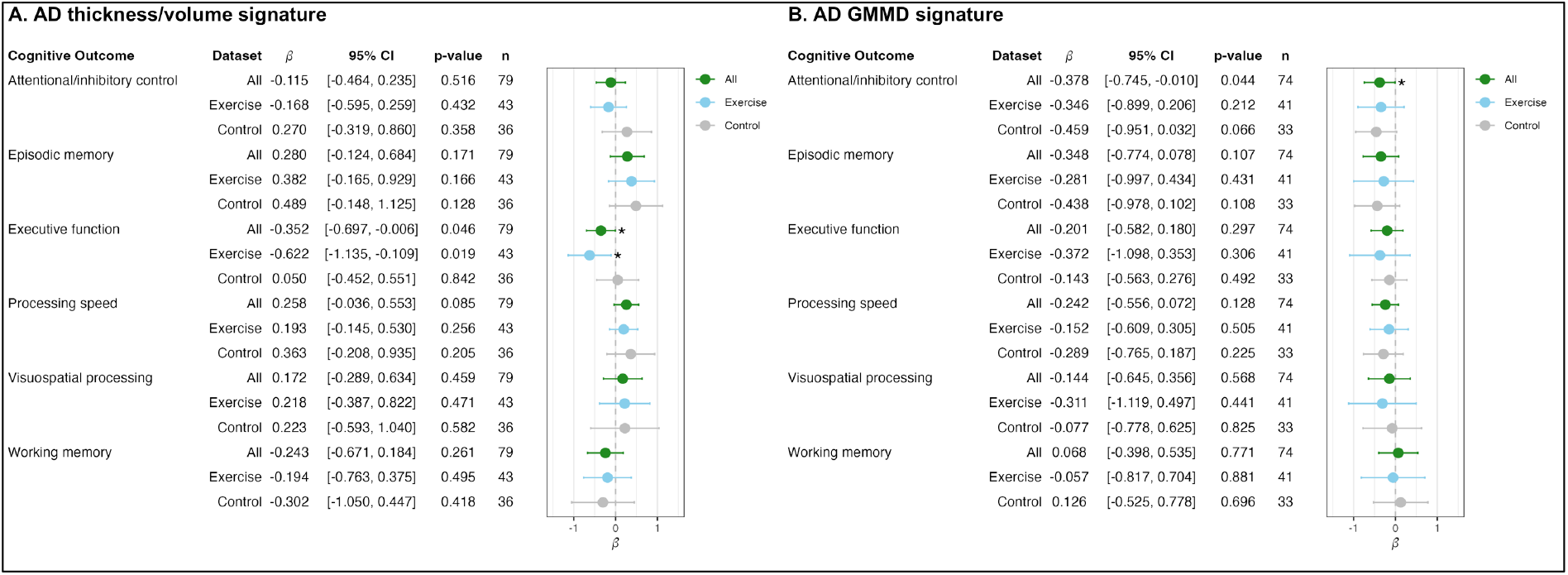
Associations between post–pre z-score differences of Alzheimer’s disease (AD) brain signatures and cognitive domains. (A) Across all participants and in the exercise group, the AD thickness/volume signature was negatively associated with changes in executive function. (B) Across all participants, AD gray matter mean diffusivity (GMMD) was negatively associated with changes in attentional/inhibitory control. Abbreviations: AD, Alzheimer’s disease; GMMD, gray matter mean diffusivity. * *p* < 0.05.

### Testing whether AD brain signatures mediate the effects of RE on cognitive outcomes

Mediation results for the AD thickness/volume signature and AD GMMD signature on cognitive outcomes are presented in Supplementary Table 6. No mediation effects were observed for either AD brain signatures.

## DISCUSSION

The study aimed to (1) examine the effects of a 24-week RE program on AD brain signatures in cognitively normal older adults; and (2) identify moderating factors, assess the association between AD brain signatures changes and cognitive outcomes, including testing whether AD brain signatures mediate an association between RE group allocation and cognitive changes. The RE group showed small reductions in the AD thickness/volume signature after 24-weeks compared to the CG, with Aβ burden moderating the effect, such that there were greater declines in Aβ-positive participants following exercise. No group differences were found in the AD GMMD signature, or moderation by any of the tested factors. Greater decreases in AD thickness/volume were associated with greater improvements in executive function. A similar pattern was found between AD GMMD signature changes and improvements in attentional/inhibitory control.

### Effect of the exercise intervention on AD brain signatures

Contrary to our expectations, we found that the RE intervention reduced the AD thickness/volume signature in cognitively healthy older adults. This finding contrasts with previous research; for example, one study reported no effect on the AD signature following a 2-year multimodal intervention in older adults at increased dementia risk.^14^ Similarly, a 12-month, multisite RCT in older adults with amnestic mild cognitive impairment found no effect on the AD signature after either moderate-to-high intensity aerobic training or lower-intensity stretching, balance, and range of motion exercises, compared to a no-intervention group.^43^ It is relevant to consider the differences in the methods used to compute AD signature in these studies, as well as the characteristics of the participants and populations, which could explain some of the variability in results. However, despite methodological differences, our sensitivity analyses using AD signatures based on cortical thickness and volumes further support the robustness of our results.

Research assessing the effects of exercise interventions on GMMD is scarce, and no studies have specifically examined the AD GMMD signature. In our study, we found no effect of the RE intervention on the AD GMMD signature. In contrast, one study reported increases in GMMD in the insula and cerebellum in both mild cognitive impairment and cognitively healthy controls after a 3-month supervised walking intervention, suggesting improvements in neural circuits and microstructural remodeling.^15^ These contrasting findings may reflect differential GMMD responses depending of the type of exercise; however, differences in study populations and outcome measures limit direct comparisons.

The observed group differences changes in the AD thickness/volume signature may reflect differential characteristic of early preclinical AD. A biphasic trajectory of macrostructural and microstructural brain changes has been proposed in the AD continuum,^44,45^ suggesting increased cortical thickness and decreased MD in AD-related regions during early preclinical AD, followed by the opposite pattern as the disease progresses and symptoms emerge. This model was supported by a study tracking longitudinal changes in AD thickness/volume and GMMD signatures over time.^46^ Similarly, another study found reduced atrophy rates in preclinical AD stage 1 compared to stage 0, followed by increased atrophy in stages 2 and 3.^47^ Thus, the RE program may have helped to prevent early preclinical AD-related changes on AD thickness/volume signature in Aβ-positive participants, whereas the CG followed the expected trajectory.

Possible explanations for the biphasic trajectory of macrostructural and microstructural brain changes include biomarker alterations and neuroinflammation. In the early stages of AD, astrocytosis has been linked to increased cortical thickness and reduced MD,^48^ and increased levels of glial fibrillary acidic protein are associated with elevated Aβ.^49^ An amyloid-triggered inflammatory response has been suggested as the cause of the increase in cortical thickness and reduction of MD during the early preclinical stage of AD stage.^44^ Moreover, some studies have reported cortical thickening in AD-related regions in Aβ-positive participants compared to Aβ-negative participants during the preclinical stage of AD.^50,51^ Considering that our Aβ-positive participants were in the early preclinical AD stages, neuroinflammation and AD-related biomarker status might partially explain the observed changes in the AD thickness/volume signature. Further research should confirm this and explore the impact of exercise on the links between neuroinflammation, AD-related biomarkers, and AD brain signatures.

### AD brain signatures and cognition

Both macro- and microstructural brain changes have been suggested as potential mechanisms underlying cognitive changes.^52^ Therefore, we explored whether changes in AD brain signatures were associated with cognitive performance.

An unexpected finding was the negative association between changes in the AD thickness/volume signature and executive function. Specifically, in the exercise group, reductions in the AD thickness/volume signature were associated with improvements in executive function. Consistent with our findings, a previous RCT reported reductions in whole brain volume accompanied by improvements in executive function after 12 months of either once-weekly or twice-weekly RE, compared to a control group (twice-weekly balance and tone training).^53^ While our findings may suggest a potential mediating role of the AD thickness/volume signature in the relationship between RE and executive function, no significant mediation effect was observed. These intriguing findings highlight the need for further research to replicate and clarify the underlying mechanisms linking exercise, AD brain signatures, and cognition.

Regarding microstructural changes, we found a negative association between the AD GMMD signature and attentional/inhibitory control. This contrasts with a previous study that reported increases in cortical MD in response to a 3-month supervised walking intervention, which were associated with improvements in verbal fluency and episodic memory in both individuals with mild cognitive impairment and cognitively healthy participants.^15^ These discrepancies may be explained by differences in exercise modality, exercise intensity, participant characteristics, or methodological approaches. Further studies are needed to confirm and better understand these associations.

### Strengths and limitations

A key strength of this study was the use of structural and diffusion brain imaging to assess exercise effects on AD brain signatures. Additionally, PET/CT imaging provided valuable information of brain Aβ deposition, considering the identified moderation effect of Aβ burden. Our findings remained significant even after sensitivity analyses excluded low-quality images. Given the variability in AD structural brain signature methodologies, we conducted sensitivity analyses using different approaches and found consistent results across methods. Furthermore, we explicitly detailed our structural and diffusion pipelines to enhance reproducibility. The exercise protocol itself is cost-effective, reproducible, and applicable to older adult populations. However, the study has limitations. It focused on cognitively normal older adults, so the results may not be generalizable to older adult populations with more severe cognitive decline. Additionally, only 21% of the sample was categorized as Aβ-positive, meaning the moderation effects of Aβ burden should be interpreted with caution. The lack of follow-up prevented assessment of potential residual effects. To fully understand changes in AD brain signatures within the biphasic model, larger interventions with extended follow-up are needed to confirm the potential of exercise for delaying early structural and diffusion changes in preclinical AD. Furthermore, studies with a larger number of Aβ-positive participants are necessary to corroborate our findings.

## CONCLUSION

The 24-week RE program led to a reduction in the AD thickness/volume signature, particularly among Aβ-positive participants. In contrast, the RE intervention did not affect the AD GMMD signature. Interestingly, reductions in the AD thickness/volume signature were associated with improvements in executive function. These findings suggest a potential beneficial effect of RE on brain health by delaying structural changes characteristic of the preclinical stages of AD. Future research should confirm our results and examine the clinical implications of these findings on cognitive function and brain aging to achieve a more comprehensive understanding of the impact of RE.

## Data Availability

All data produced in the present study are available upon reasonable request to the authors.

## ACKNOWLEDGEMENTS

The authors would like to thank the participants who took part in this study. Additionally, we acknowledge María Teresa Rodríguez Palacios for her contribution to the visual quality control of diffusion-weighted images.

