## Supplementary material for "Effects of a 24-week resistance exercise program on Alzheimer’s disease brain signatures in cognitively normal older adults: results from the AGUEDA trial"

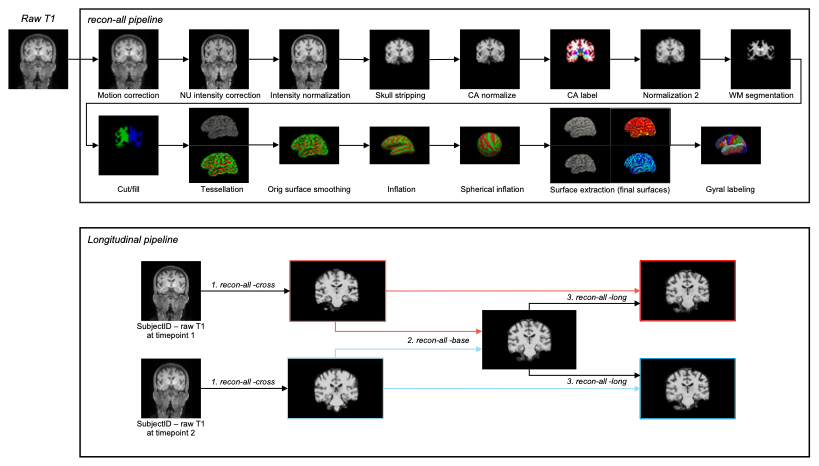
**Supplementary Figure 1.** Longitudinal processing pipeline using FreeSurfers’s recon-all. Abbreviation: WM, white matter.

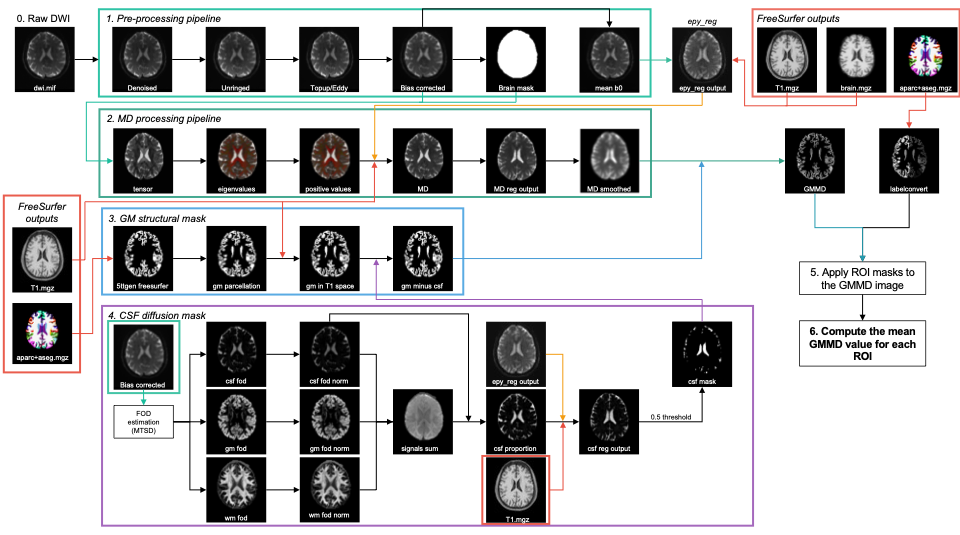
**Supplementary Figure 2.** Processing pipeline for gray matter mean diffusivity estimation. Abbreviations: csf, cerebrospinal fluid; dwi, diffusion-weighted image; gm, gray matter; GMMD, gray matter mean diffusivity; MD, mean diffusivity; ROI, region of interest; wm, white matter.

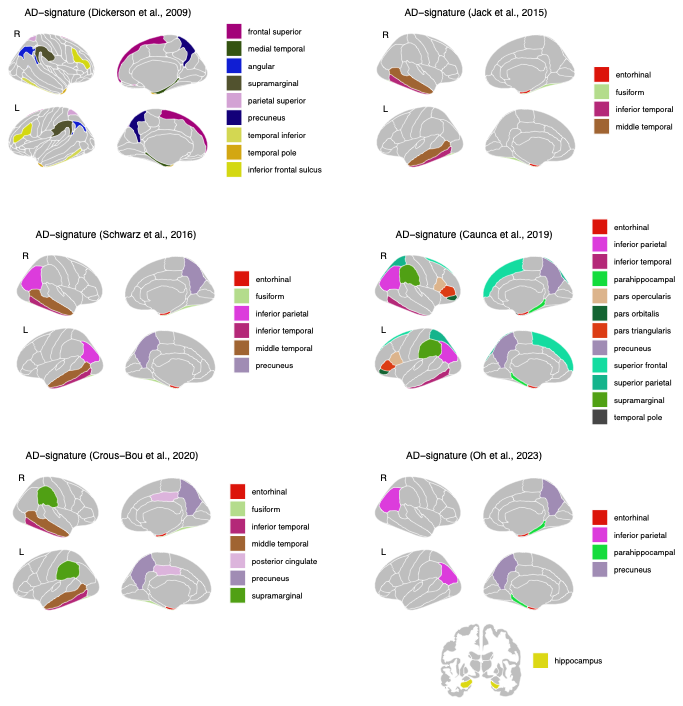

**Supplementary Figure 3.** Brain regions used by the methodologies included to compute Alzheimer’s disease (AD) signatures. Most AD-signatures were derived by averaging individual cortical thicknesses across bilateral regions of interest (ROIs) and then z-scored; however, Oh et al. (2023) utilized brain volumes. ROIs for the Jack et al. (2015), Schwarz et al. (2016), Caunca et al. (2019), Crous-Bou et al. (2020), and Oh et al. (2023) approaches were obtained from FreeSurfer's outputs (‘lh.aparc.stats’ and ‘rh.aparc.stats’ files), which include the Desikan-Killiany atlas parcellation (Desikan et al., 2006). Additionally, hippocampal volumes for the Oh et al. (2023) approach were obtained from the ‘aseg.stats’ file (a FreeSurfer output), which also contains the Desikan-Killiany atlas parcellation. ROIs for the Dickerson et al. (2009) approach were obtained from FreeSurfer’s outputs (‘lh.aparc.a2009s.stats’ and ‘rh.aparc.a2009s.stats’ files), which include the Destrieux atlas parcellation (Destrieux et al., 2009).

| Supplementary Table 1. Consolidated Standards of Reporting Trials checklist. | | | |
| --- | --- | --- | --- |
| Section/topic | No | CONSORT 2025 checklist item description | Reported on page no. |
| **Title and abstract** | | |  |
| Title and structured abstract | 1a | Identification as a randomised trial | 1 |
|  | 1b | Structured summary of the trial design, methods, results, and conclusions | 1 |
| **Open science** | | |  |
| Trial registration | 2 | Name of trial registry, identifying number (with URL) and date of registration | 2 |
| Protocol and statistical analysis plan | 3 | Where the trial protocol and statistical analysis plan can be accessed | 2, Ref 16 |
| Data sharing | 4 | Where and how the individual de-identified participant data (including data dictionary), statistical code and any other materials can be accessed | 2 |
| Funding and conflicts of interest | 5a | Sources of funding and other support (eg, supply of drugs), and role of funders in the design, conduct, analysis and reporting of the trial | 11 |
|  | 5b | Financial and other conflicts of interest of the manuscript authors | 11 |
| **Introduction** | | |  |
| Background and rationale | 6 | Scientific background and rationale | 2 |
| Objectives | 7 | Specific objectives related to benefits and harms | 2 |
| **Methods** | | |  |
| Patient and public involvement | 8 | Details of patient or public involvement in the design, conduct and reporting of the trial | 2 |
| Trial design | 9 | Description of trial design including type of trial (eg, parallel group, crossover), allocation ratio, and framework (eg, superiority, equivalence, non-inferiority, exploratory) | 2, Ref 16 |
| Changes to trial protocol | 10 | Important changes to the trial after it commenced including any outcomes or analyses that were not prespecified, with reason | Ref 17 |
| Trial setting | 11 | Settings (eg, community, hospital) and locations (eg, countries, sites) where the trial was conducted | 2 |
| Eligibility criteria | 12a | Eligibility criteria for participants | 2, Ref 16 |
|  | 12b | If applicable, eligibility criteria for sites and for individuals delivering the interventions (eg, surgeons, physiotherapists) | NA |
| Intervention and comparator | 13 | Intervention and comparator with sufficient details to allow replication. If relevant, where additional materials describing the intervention and comparator (eg, intervention manual) can be accessed | 2, Ref 19 |
| Outcomes | 14 | Prespecified primary and secondary outcomes, including the specific measurement variable (eg, systolic blood pressure), analysis metric (eg, change from baseline, final value, time to event), method of aggregation (eg, median, proportion), and time point for each outcome | 4 |
| Harms | 15 | How harms were defined and assessed (eg, systematically, non-systematically) | NA |
| Sample size | 16a | How sample size was determined, including all assumptions supporting the sample size calculation | 2, Ref 16 |
|  | 16b | Explanation of any interim analyses and stopping guidelines | NA |
| Randomisation: |  |  |  |
| Sequence generation | 17a | Who generated the random allocation sequence and the method used | 2, Ref 16 |
|  | 17b | Type of randomisation and details of any restriction (eg, stratification, blocking and block size) | 2, Ref 16 |
|  |  |  | **Reported on page no.** |
| Allocation concealment mechanism | 18 | Mechanism used to implement the random allocation sequence (eg, central computer/telephone; sequentially numbered, opaque, sealed containers), describing any steps to conceal the sequence until interventions were assigned | 2, Ref 16 |
| Implementation | 19 | Whether the personnel who enrolled and those who assigned participants to the interventions had access to the random allocation sequence | 2, Ref 16 |
| Blinding | 20a | Who was blinded after assignment to interventions (eg, participants, care providers, outcome assessors, data analysts) | 2, Ref 16 |
|  | 20b | If blinded, how blinding was achieved and description of the similarity of interventions | 2, Ref 16 |
| Statistical methods | 21a | Statistical methods used to compare groups for primary and secondary outcomes, including harms | 5 |
|  | 21b | Definition of who is included in each analysis (eg, all randomised participants), and in which group | 5, 6, Table 1, Suppl Table 3 |
|  | 21c | How missing data were handled in the analysis | 5 |
|  | 21d | Methods for any additional analyses (eg, subgroup and sensitivity analyses), distinguishing prespecified from post hoc | 5 |
| **Results** | | |  |
| Participant flow, including flow diagram | 22a | For each group, the numbers of participants who were randomly assigned, received intended intervention, and were analysed for the primary outcome | 2, Table 1, Suppl Table 3, Suppl Table 4 |
|  | 22b | For each group, losses and exclusions after randomisation, together with reasons | Ref 17 |
| Recruitment | 23a | Dates defining the periods of recruitment and follow-up for outcomes of benefits and harms | 2 |
|  | 23b | If relevant, why the trial ended or was stopped | NA |
| Intervention and comparator delivery | 24a | Intervention and comparator as they were actually administered (eg, where appropriate, who delivered the intervention/comparator, how participants adhered, whether they were delivered as intended (fidelity)) | Ref 17 |
|  | 24b | Concomitant care received during the trial for each group | Ref 17 |
| Baseline data | 25 | A table showing baseline demographic and clinical characteristics for each group | 6 |
| Numbers analysed,  outcomes and estimation | 26 | For each primary and secondary outcome, by group:  ● the number of participants included in the analysis  ● the number of participants with available data at the outcome time point  ● result for each group, and the estimated effect size and its precision (such as 95% confidence interval)  ● for binary outcomes, presentation of both absolute and relative effect size | 6, 7, 8, 9, Table 1, Suppl Table 3 |
| Harms | 27 | All harms or unintended events in each group | Ref 17 |
| Ancillary analyses | 28 | Any other analyses performed, including subgroup and sensitivity analyses, distinguishing pre-specified from post hoc | 6, 7, Suppl Table 4, Suppl Table 5 |
| **Discussion** | | |  |
| Interpretation | 29 | Interpretation consistent with results, balancing benefits and harms, and considering other relevant evidence | 9, 10 |
| Limitations | 30 | Trial limitations, addressing sources of potential bias, imprecision, generalisability, and, if relevant, multiplicity of analyses | 10 |

| **Supplementary Table 2**. Cognitive domains, and their corresponding cognitive tests. | |
| --- | --- |
| Domain | Cognitive test |
| Attentional/inhibitory control | Dimensional Change Card Sort Task |
|  | Flanker |
|  | Stroop Task (incongruent trial) |
|  | Trail Making Test (part B) |
| Episodic memory | Montreal Cognitive Assessment (delayed recall) |
|  | Picture Sequence Memory Test |
|  | Rey Auditory Verbal Learning Test |
|  | Rey-Osterrieth Complex Figure Test |
| Executive function | Digit Symbol Substitution Test |
|  | Dimensional Change Card Sort Test |
|  | Trail Making Test |
|  | Spatial Working Memory Test |
| Processing speed | Digit Symbol Substitution Test |
|  | Trail Making Test (part A) |
| Visuospatial processing | Montreal Cognitive Assessment (clock drawing) |
|  | Wechsler Adult Intelligence Scale (matrix reasoning and block design) |
| Working memory | N-Back Working Memory Task |
|  | List Sorting Working Memory Test |
|  | Spatial Working Memory Task |

| **Supplementary Table 3.** Inclusion and exclusion of participants considering brain image quality for main and sensitivity analyses by AD brain signature. | | | |
| --- | --- | --- | --- |
| *For AD thickness/volume signature analyses* | Pre | Post | All |
| All T1 images | 90 | 79 | 170 |
| **Included for main analysis** | **90** | **79** | **170** |
| Moderate QC rating with issues in parcellation of ROIs | 11 | 10 | 21 |
| **Included for sensitive analysis** | **79** | **69** | **149** |
| *For AD GMMD signature analyses* |  |  |  |
| All DWI | 90 | 79 | 169 |
| Incorrect phase direction | 3 | 0 | 3 |
| Visual DWI QC rated as severe | 0 | 1 | 1 |
| Automatic QC ≥ 2 exceeded thresholds | 1 | 1^a^ | 2 |
| **Included for main analysis** | **86** | **78** | **164** |
| Parcellation issues of ROIs in the FreeSurfer output | 7 | 10 | 17 |
| Visual DWI QC rated as moderate | 5^b^ | 8 | 13 |
| **Included for sensitive analysis** | **75** | **60** | **135** |
| Abbreviations: AD, Alzheimer’s disease; DWI, diffusion-weighted images; GMMD, gray matter mean diffusivity; ROIs, regions of interest; QC, quality control.  ^a^ Corresponded to a participant image rated as severe in the visual DWI QC.  ^b^ Corresponded to a participant image with parcellation issues in the FreeSurfer output. | | | |

| **Supplementary Table 4**. Estimated marginal means in Alzheimer’s disease signature cortical thickness or volume using additional methodologies, and results of the sensitivity analysis excluding images with parcellation issues, stratified by amyloid beta status. | | | | | | | | | | | |
| --- | --- | --- | --- | --- | --- | --- | --- | --- | --- | --- | --- |
|  | Main analysis | | | | |  | Sensitivity analysis | | | | |
|  | **Pre** | **Post** | | |  |  | **Pre** | **Post** | | |  |
|  | **All** | **RE** | **CG** | **Group difference** | **p-value** |  | **All** | **RE** | **CG** | **Group difference** | **p-value** |
| **Overall** | n = 90 | n = 46 | n = 44 |  |  |  | n = 79 | n = 41 | n = 38 |  |  |
| AD thickness/volume signature | 0 [-0.21;0.21] | -0.07 [-0.31;0.17] | 0.15 [-0.09;0.40] | -0.23 [-0.43;0.02] | **0.032** |  | 0 [-0.22;0.22] | -0.03 [-0.27;0.22] | 0.17 [-0.08;0.43] | -0.20 [-0.42;0.02] | 0.070 |
| AD signature (Dickerson et al., 2009) | 0 [-0.21;0.21] | -0.22 [-0.46;0.02] | 0.08 [-0.16;0.33] | -0.30 [-0.52;-0.09] | **0.007** |  | 0 [-0.22;0.22] | -0.23 [-0.50;0.04] | 0.06 [-0.22;0.34] | -0.29 [-0.54;-0.04] | **0.025** |
| AD signature (Jack et al., 2015) | 0 [-0.21;0.21] | -0.15 [-0.38;0.09] | 0.12 [-0.12;0.36] | -0.27 [-0.48;-0.06] | **0.012** |  | 0 [-0.22;0.22] | -0.20 [-0.46;0.06] | 0.09 [-0.19;0.36] | -0.28 [-0.53;-0.04] | **0.025** |
| AD signature (Schwarz et al., 2016) | 0 [-0.21;0.21] | -0.19 [-0.43;0.05] | 0.13 [-0.12;0.38] | -0.32 [-0.54;-0.10] | **0.005** |  | 0 [-0.22;0.22] | -0.24 [-0.50;0.03] | 0.10 [-0.18;0.37] | -0.33 [-0.59;-0.07] | **0.014** |
| AD signature (Caunca et al., 2019) | 0 [-0.21;0.21] | -0.18 [-0.42;0.05] | 0.05 [-0.19;0.29] | -0.24 [-0.44;-0.04] | **0.020** |  | 0 [-0.22;0.22] | -0.20 [-0.46;0.05] | 0.02 [-0.24;0.29] | -0.23 [-0.46;0.00] | 0.054 |
| AD signature (Crous-Bou et al., 2020) | 0 [-0.21;0.21] | -0.17 [-0.40;0.07] | 0.10 [-0.14;0.35] | -0.27 [-0.48;-0.06] | **0.011** |  | 0 [-0.22;0.22] | -0.21 [-0.46;0.05] | 0.07 [-0.19;0.34] | -0.28 [-0.52;-0.04] | **0.020** |
| AD signature^a^ (Oh et al., 2023) | 0 [-0.21;0.21] | -0.15 [-0.37;0.07] | 0.03 [-0.20;0.25] | -0.18 [-0.30;-0.05] | **0.007** |  | 0 [-0.22;0.22] | -0.16 [-0.40;0.08] | 0.01 [-0.23;0.25] | -0.17 [-0.32;-0.03] | **0.022** |
| **Aβ-negative older adults** | n = 71 | n = 38 | n = 33 |  |  |  | n = 64 | n = 35 | n = 29 |  |  |
| AD thickness/volume signature | 0.02 [-0.22;0.27] | -0.01 [-0.27;0.26] | 0.09 [-0.19;0.38] | -0.10 [-0.33;0.13] | 0.394 |  | 0 [-0.26;0.26] | 0.01 [-0.27;0.29] | 0.09 [-0.21;0.39] | -0.08 [-0.33;0.17] | 0.519 |
| AD signature (Dickerson et al., 2009) | 0.06 [-0.18;0.30] | -0.11 [-0.38;0.17] | 0.05 [-0.24;0.33] | -0.15 [-0.38;0.08] | 0.192 |  | 0.03 [-0.22;0.28] | -0.13 [-0.43;0.16] | -0.02 [-0.33;0.30] | -0.12 [-0.37;0.14] | 0.378 |
| AD signature (Jack et al., 2015) | 0.01 [-0.23;0.26] | -0.10 [-0.38;0.17] | 0.09 [-0.20;0.38] | -0.20 [-0.43;0.04] | 0.099 |  | -0.01 [-0.27;0.26] | -0.15 [-0.45;0.15] | 0.05 [-0.27;0.37] | -0.20 [-0.48;0.08] | 0.157 |
| AD signature (Schwarz et al., 2016) | 0.03 [-0.21;0.27] | -0.12 [-0.40;0.16] | 0.09 [-0.20;0.38] | -0.21 [-0.45;0.03] | 0.082 |  | 0 [-0.25;0.26] | -0.16 [-0.46;0.14] | 0.04 [-0.28;0.37] | -0.21 [-0.48;0.07] | 0.144 |
| AD signature (Caunca et al., 2019) | 0.02 [-0.23;0.27] | -0.12 [-0.39;0.15] | -0.01 [-0.29;0.27] | -0.11 [-0.33;0.11] | 0.335 |  | -0.01 [-0.27;0.25] | -0.16 [-0.45;0.13] | -0.08 [-0.39;0.23] | -0.07 [-0.33;0.18] | 0.556 |
| AD signature (Crous-Bou et al., 2020) | 0.03 [-0.21;0.28] | -0.10 [-0.38;0.17] | 0.08 [-0.21;0.36] | -0.18 [-0.41;0.05] | 0.121 |  | 0 [-0.26;0.26] | -0.15 [-0.44;0.15] | 0.02 [-0.29;0.34] | -0.17 [-0.43;0.09] | 0.199 |
| AD signature^a^ (Oh et al., 2023) | -0.02 [-0.26;0.21] | -0.13 [-0.37;0.11] | -0.05 [-0.30;0.19] | -0.07 [-0.20;0.06] | 0.273 |  | -0.03 [-0.26;0.21] | -0.12 [-0.37;0.12] | -0.08 [-0.32;0.17] | -0.05 [-0.20;0.10] | 0.512 |
| **Aβ-positive older adults** | n = 9 | n = 8 | n = 11 |  |  |  | n = 15 | n = 6 | n = 9 |  |  |
| AD thickness/volume signature | -0.08 [-0.52;0.36] | -0.35 [-0.89;0.18] | 0.28 [-0.22;0.79] | -0.64 [-1.09;-0.18] | **0.010** |  | 0 [-0.45;0.45] | -0.29 [-0.81;0.22] | 0.41 [-0.06;0.87] | -0.70 [-1.17;-0.23] | **0.007** |
| AD signature (Dickerson et al., 2009) | -0.23 [-0.65;0.20] | -0.72 [-1.24;-0.19] | 0.14 [-0.34;0.61] | -0.85 [-1.38;-0.33] | **0.004** |  | -0.13 [-0.65;0.40] | -0.77 [-1.43;-0.11] | 0.25 [-0.31;0.82] | -1.02 [-1.67;-0.37] | **0.006** |
| AD signature (Jack et al., 2015) | -0.06 [-0.46;0.35] | -0.38 [-0.84;0.08] | 0.20 [-0.21;0.62] | -0.59 [-1.06;-0.11] | **0.018** |  | 0.03 [-0.42;0.47] | -0.48 [-1.01;0.05] | 0.20 [-0.26;0.66] | -0.68 [-1.25;-0.11] | **0.023** |
| AD signature (Schwarz et al., 2016) | -0.11 [-0.54;0.32] | -0.54 [-1.04;-0.04] | 0.21 [-0.24;0.66] | -0.75 [-1.29;-0.22] | **0.009** |  | -0.02 [-0.54;0.50] | -0.66 [-1.29;-0.03] | 0.25 [-0.28;0.79] | -0.91 [-1.59;-0.23] | **0.013** |
| AD signature (Caunca et al., 2019) | -0.07 [-0.48;0.34] | -0.49 [-0.97;-0.01] | 0.20 [-0.24;0.65] | -0.69 [-1.14;-0.24] | **0.005** |  | 0.06 [-0.40;0.52] | -0.49 [-1.01;0.03] | 0.36 [-0.10;0.81] | -0.85 [-1.35;-0.34] | **0.004** |
| AD signature (Crous-Bou et al., 2020) | -0.12 [-0.53;0.29] | -0.45 [-0.90;0.00] | 0.16 [-0.24;0.57] | -0.62 [-1.08;-0.16] | **0.012** |  | 0 [-0.46;0.46] | -0.56 [-1.06;-0.06] | 0.23 [-0.21;0.67] | -0.79 [-1.28;-0.30] | **0.005** |
| AD signature^a^ (Oh et al., 2023) | 0.09 [-0.42;0.60] | -0.31 [-0.89;0.27] | 0.28 [-0.29;0.84] | -0.59 [-0.91;-0.26] | **0.002** |  | 0.11 [-0.56;0.78] | -0.44 [-1.20;0.32] | 0.31 [-0.44;1.05] | -0.75 [-1.10;-0.40] | **0.001** |
| Abbreviations: Aβ, amyloid beta; AD, Alzheimer’s disease; CG, wait-list control group; RE, resistance exercise group. Data are presented as mean change [95% confidence interval]. ^a^ Adjusted for intracranial volume. All AD signatures were computed using cortical thickness, except for Oh et al. (2023), which utilized brain volumes. | | | | | | | | | | | |

| **Supplementary Table 5**. Estimated marginal means in Alzheimer’s disease signature gray matter mean diffusivity, and results of the sensitivity analysis excluding low-quality images | | | | | | | | | | | |
| --- | --- | --- | --- | --- | --- | --- | --- | --- | --- | --- | --- |
|  | Main analysis | | | | |  | Sensitivity analysis | | | | |
|  | **Pre** | **Post** | | |  |  | **Pre** | **Post** | | |  |
|  | **All** | **RE** | **CG** | **Group difference** | **p-value** |  | **All** | **RE** | **CG** | **Group difference** | **p-value** |
|  | n = 89 | n = 46 | n = 43 |  |  |  | n = 77 | n = 40 | n = 37 |  |  |
| AD GMMD signature | -0.01 [-0.21;0.20] | 0.05 [-0.20;0.30] | -0.03 [-0.28;0.23] | -0.8 [-0.13;0.29] | 0.457 |  | -0.01 [-0.24;0.22] | 0.07 [-0.21;0.36] | 0.01 [-0.29;0.3] | 0.07 [-0.2;0.33] | 0.625 |
| Abbreviations: AD, Alzheimer’s disease; CG, wait-list control group; GMMD, gray matter mean diffusivity; RE, resistance exercise group. Data are presented as mean change [95% confidence interval]. | | | | | | | | | | | |

| **Supplementary Table 6**. Mediation analysis results. | | | | | | |
| --- | --- | --- | --- | --- | --- | --- |
| Mediator | Outcome | Path a  (X 🡪 M) | Path b  (M 🡪 Y) | Indirect Effect  (a×b) [95%CI] | Direct Effect (c') | Total Effect (c) |
| AD thickness/volume signature | Attentional/inhibitory control | **-0.213** | 0.119 | -0.026 [-0.136; 0.049] | **0.502** | **0.475** |
|  | Episodic memory | **-0.213** | 0.38 | -0.08 [-0.257; 0.02] | **0.466** | **0.386** |
|  | Executive function | **-0.213** | -0.301 | 0.064 [-0.015; 0.188] | 0.133 | 0.197 |
|  | Processing speed | **-0.213** | 0.263 | -0.053 [-0.167; 0.017] | 0.047 | -0.006 |
|  | Visuospatial processing | **-0.213** | 0.276 | -0.058 [-0.208; 0.04] | 0.103 | 0.046 |
|  | Working memory | **-0.213** | -0.04 | 0.008 [-0.083; 0.114] | 0.02 | 0.028 |
| AD GMMD signature | Attentional/inhibitory control | 0.073 | **-0.447** | -0.032 [-0.142; 0.058] | **0.509** | **0.477** |
|  | Episodic memory | 0.073 | -0.38 | -0.028 [-0.143; 0.066] | **0.413** | **0.385** |
|  | Executive function | 0.073 | -0.254 | -0.019 [-0.098; 0.04] | 0.222 | 0.204 |
|  | Processing speed | 0.073 | -0.273 | -0.018 [-0.097; 0.049] | 0.02 | 0.002 |
|  | Visuospatial processing | 0.073 | -0.251 | -0.019 [-0.107; 0.042] | 0.072 | 0.053 |
|  | Working memory | 0.073 | -0.025 | -0.002 [-0.06; 0.052] | 0.029 | 0.027 |
| Abbreviations: AD, Alzheimer’s disease; GMMD, gray matter mean diffusivity. Significant results shown in bold (p <0.05). | | | | | | |
